# Altruistic punishment supports the persistence of social norms for infectious diseases

**DOI:** 10.64898/2026.07.30.26358890

**Authors:** Sefah Frimpong, Chris T. Bauch

## Abstract

In the face of an epidemic where a population behaviour both influences disease transmission and reacts to it, social processes can generate norms to support socially beneficial behaviour. Most mathematical models of coupled behaviour–disease dynamics treat norms as pre-existing rather than explaining how they are maintained. Here, we investigate whether altruistic punishment can sustain a social distancing norm when individuals may defect, cooperate without punishing, or cooperate while paying a cost to punish defectors. We couple a transmission model to an imitation model for these three strategies. Disease prevalence affects behavioural payoffs, while the behavioural composition modifies transmission. We also compare this coupled system with a control where behavioural decisions respond to a fixed prevalence. We find a wide parameter regime corresponding to the establishment of an injunctive social norm in support of social distancing, where the punisher strategy is widespread. Persistence may occur through stable states where punishers or dominant. Disease–behaviour feedback can also create oscillations (where the three strategies succeed one another in response to epidemic waves) or tipping points (sharp transitions between all-defector and cooperative states). These effects do not occur in the uncoupled model, although there are still broad parameter regimes where a social norm persists. Our findings show that costly peer punishment can support persistence of social norms that mitigate disease transmission. More broadly, endogenous epidemic feedback can qualitatively change the conditions under which cooperation and punishment are sustained, producing tipping points and long-term behavioural–epidemiological cycles that fixed-payoff models cannot capture.

**Significance Statement:** Populations both shape disease transmission and respond to prevalence, allowing social processes to sustain norms. We examine whether altruistic punishment can maintain social distancing where individuals may defect, cooperate, or cooperate while also paying to punish defectors. A transmission model is coupled to imitation dynamics, so disease prevalence and behaviour influence one another. We compare this system with a control based on fixed perceived prevalence. Across broad parameter ranges, punishers persist and support a social distancing norm. Coupled feedback can also generate oscillations, as strategies rise and fall with epidemic waves, and tipping points between defection and cooperation. These features are absent from the uncoupled model. Thus, epidemic feedback can alter how cooperation, punishment, and disease control norms persist.

## Introduction

Social norms are pervasive and powerful, governing a wide range of phenomena^1–5^. This includes cooperation in public goods problems^6^, in which members of a group face a dilemma: all members benefit equally from a public good that requires contributions from members to maintain. As a result, there is an incentive for members to ‘free-ride’ by enjoying benefits from those who contribute while not paying for any contributions^7,8^. This gives rise to a game where members must decide their strategy: cooperate or defect^9^.

Free-riding in game theoretical models can prevent outcomes that are best for the group as a whole^10^. However, in real populations, humans punish defectors even in one-shot interactions^11^ and in populations of genetically unrelated individuals^12^. This punishing behaviour can sustain cooperation by imposing a penalty for non-cooperation, such as through fines or ostracism. If the punisher must pay some cost to punish defectors, then the behaviour can be considered altruistic punishment, since the act of punishing becomes an altruistic act if it requires a cost to impose. Altruistic punishment can support cooperation in public goods problems under the right conditions^13–15^. Altruistic punishment can take several forms, such as defined penalties for free-riders, emotional responses—anger or indignation towards defectors, and the restriction of defectors from accessing other social facilities–schools, public places, and, in extreme cases, imprisonment^8^. Punishing defectors achieves two main outcomes: it serves as a deterrent that discourages potential free-riders, and it forces existing free-riders to change their behaviour to cooperate^16^.

When punishing imposes a cost on the punisher, it becomes possible to distinguish ‘cooperators’ (who cooperate by contributing to maintaining the public good but do not punish) from ‘altruistic punishers’ (who both cooperate and punish). However, if punishing defectors imposes a cost on the punisher, why do they do it? In these situations, it could be said that cooperators are second-order defectors who are willing to cooperate but are not willing to pay the cost of punishing defectors^17^. Specific conditions are required to allow altruistic punishment to emerge and be maintained, thereby sustaining the public good, despite its higher cost to the individual. Previous work has studied the role of factors such as the cost of punishing^18^, population size^14^, multi-level selection^19,20^, and individual and group heterogeneity^21^. The role of punishment has also been studied in the context of coupled human-and-natural systems^22^, where groups must make decisions about the sustainable harvesting of limited natural resources. A particular focus is on the conditions under which norms about harvesting can be stable and produce sustainable outcomes^1,23^. A population where altruistic punishment of defectors through mechanisms such as sanctions is stable can be said to exemplify a social norm^12,24,25^, particularly an injunctive social norm^26^.

Human behaviour (and especially processes like social learning and social norms) is also integral to infectious diseases^27^, which have long posed a significant threat to human well-being^28,29^. It is perhaps not surprising that infectious diseases and social processes are inextricably bound, since our social connections overlap with many routes of disease transmission. As a result, it is perhaps not surprising that norms concerning infectious diseases are widespread, covering diverse aspects of human behaviour^30,31^. Evidence suggests that disgust, which predates human culture, may serve as a motivator for disease avoidance^32,33^. As a result, it is easy to see how the emergence of social norms around hygiene and contact precautions could be supported by this^33^.

The control of infectious diseases through adherence to pharmaceutical or non-pharmaceutical measures can be considered a public goods problem. Contact precautions reduce the prevalence of infection in the entire population, thus benefiting not just the individuals practicing the measures but also the rest of the population. However, these measures can be costly, and some individuals ‘defect’ by choosing not to practice them. Hence, social norms that support contact precautions and other pharmaceutical and non-pharmaceutical measures can become strong in the face of infection risks to a population^30,34–36^, helping to mitigate the spread of infectious diseases by reducing contact between infectious and susceptible individuals^37–40^. In situations of high infection risk, social norms that support contact precautions become very strong relative to other cultural or meta norms, as the population seeks to protect itself^41–43^. In such instances, compliance becomes strict and punishable. In contrast, social distancing norms in an environment of ebbing disease prevalence are weaker^42^. Thus, infectious disease dynamics play a role in shaping the temporal evolution of social distancing norms.

The interaction between social norm emergence and infectious disease dynamics suggests a role for a coupled behaviour-disease dynamics framework. Coupled behaviour-disease models study the interplay between the dynamics of disease outbreaks and changes in the behaviour of the population, including the adoption of control measures such as contact precautions or vaccines^44–47^. The endogenous establishment and persistence of norms have been studied in other public goods games^48,49^, but not so much in the coupled behaviour-disease literature, where social norms are typically assumed to exist *a priori*^50,51^. Because feedback can change the dynamics of cooperative behaviour, norm emergence in infectious diseases cannot be considered a special case of existing public goods models. Here, we combine insights from the literature on altruistic punishment and the literature on coupled behaviour-disease systems to determine the role of infectious diseases in the emergence and maintenance of social norms regarding infectious disease. We seek to address the questions: How do norms about infectious diseases emerge? Why are they sustained in the presence of second-order free-riding? And what role do infectious disease dynamics play in shaping them. We model social distancing as behavioural strategies where individuals can be defectors (those who want to benefit from mitigation without socially distancing themselves), cooperators (those who socially distance), and punishers (those who socially distance and punish defectors). Punishers deter defectors by imposing a penalty on free-riders (defectors) to enforce cooperation^14,52^. Infectious disease dynamics are governed by a compartmental model^53^. We characterize the dynamical regimes of the model and study their response to changes in parameter values. Modelling the cultural evolution of norms can be useful, among other reasons, because it allows exploring counter-factual worlds in which hypothesized mechanisms can be removed^2^. Hence, we also explore the role of infectious disease dynamics by comparing our model outputs to reduced systems where infection prevalence is held at fixed levels.

## Results

### Model Framework

We consider a coupled behaviour-disease model for an acute respiratory infectious disease. The model consists of a Susceptible-Infectious-Recovered (SIR) compartmental model with birth/death dynamics, coupled to imitation (replicator) dynamic equations for social distancing behaviour (see Methods for details, including model structure and parameterisation). The population is grouped into three compartments for disease progression: susceptible (S), infected (I), and recovered (R). The susceptible (S) proportion consists of individuals who have not yet been infected; the infected (I) proportion includes individuals who have come into contact with the disease causing pathogen and have become infected and infectious; the recovered (R) proportion includes individuals who have either recovered to a state of natural immunity or have died as a result of the infection. Mass action between susceptible and infected persons causes transmission of the pathogen.

Social distancing is partly effective in preventing infection. However, individuals may or may not practice it, depending on disease prevalence, the costs of social distancing, the costs of infection, the costs of being punished for not supporting the public good by practicing it, and the costs of applying that punishment. We consider three strategies: defectors (D), cooperators (C), and punishers (P). Defectors do not socially distance themselves but may pay a cost due to being punished by punishers. Cooperators practice social distancing but do not punish defectors. Punishers practice social distancing and also punish defectors at a cost to themselves. Defectors are therefore first degree free-riders, and cooperators are second degree free-riders. However, we assume that ostracism is one of the tools available to punish defectors, and thus we allow the efficacy of social distancing to be higher for punishers than for cooperators. The behaviour model is constructed using the expected payoffs for each strategy, dependent on disease prevalence. Using these payoffs, we model the rate of change of the proportion of the population adopting any of the behaviour strategies using imitation dynamics. This forms the coupled behaviour-disease model, accounting for the feedback mechanism between the two dynamical systems – disease dynamics and social distancing dynamics.

In order to study the role of temporally evolving disease dynamics, we also consider a control experiment where prevalence is kept fixed, resulting in an ‘uncoupled model’. This corresponds to the baseline assumption of many public goods games, that resource levels do not fluctuate in response to player decisions.

### Emergence of a social distancing norm

The coupled model is capable of a rich variety of dynamical behaviour (Figure 1). This includes outcomes where punishers are able to persist (with or without the presence of defectors and/or cooperators). We interpret such outcomes as representing the emergence and establishment of an injunctive social norm in support of social distancing. Norm emergence occurs for sufficiently low costs of punishment (Figure 1C). In this regime, the social distancing norm can persist through stable limit cycles, in which a temporal succession of dynamics is observed (Figure 1D,E). When disease prevalence is low and there is less incentive to practice social distancing, defection spreads and the punisher strategy declines. But this allows the virus to return, and thus the defectors decline due to the impacts of the virus, allowing punishers to resurge. Cooperators are relatively stable at this point in the parameter plane but tend to rise and fall with the defectors. In contrast, when the cost of punishment is high, punishers cannot persist (and, perhaps more surprisingly, neither can cooperators), and the population converges to an equilibrium of all defectors (Figure 1B,C). However, in this regime, the population cycles through the three strategies before eventually converging to the all-defect equilibrium (Figure 1B). If the social learning rate is too small, the population is slow to move away from the initial conditions (Figure 1A). A social distancing norm can also persist through stable co-existence between defectors and punishers, instead of limit cycles. When the cost of punishment is very low, we observe that defectors are absent, and both cooperators and punishers may co-exist at a stable nonzero level, instead of oscillating (Figure 1C). In this regime, punishers do not pay a cost to practice their strategy since the defect strategy has been eliminated from the population.

**Figure 1.**
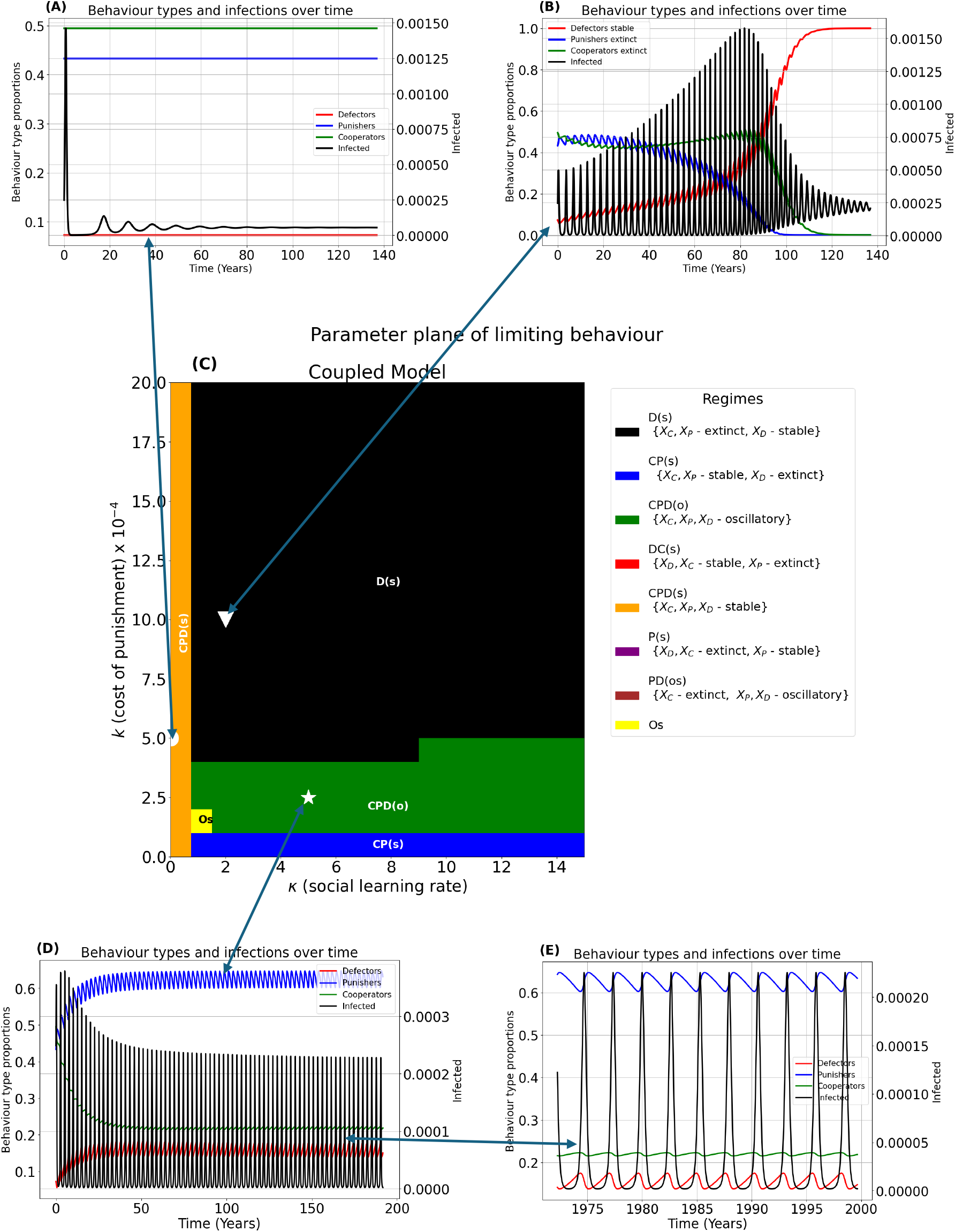
Baseline scenario with limiting behaviours for coupled model. Parameter planes for cost of punishment (*k*) and social learning rate (*κ*) show diverse limiting behaviours (**(C)**). Individual time series of points from each dynamical regime and infections are simulated a for illustration purposes (**(A),(B),(D)**).The first 200 years (**(D)**) and the corresponding last 15 years (**(E)**) are also shown. Baseline parameter values corresponding to Figure 1D appear in Table 2. Abbreviations for parameter regimes appear in Table 1.

More broadly, across both coupled and uncoupled models, we see regimes where the punisher strategy can persist, either through limit cycles or through stable coexistence (Figures 2-4). For instance, in the coupled model, the punisher strategy tends to be supported for a higher cost of the penalty or a lower cost of social distancing (Figure 2A,C,E) and the same is true for the uncoupled model (Figure 2B,D,F). However, in the coupled model, the parameter regimes of all-defect and a cooperate-punish stable state are often separated by a region where all three strategies can coexist through limit cycles (or sometimes, just punish and defect will coexist). These regimes occur for intermediate values of the cost of social distancing, the cost to punish, and the penalty for being punished.

**Table 1.** Dynamical regimes based on limiting behaviours of the coupled and uncoupled models illustrated in figures (1 - 4).

| Label | Asymptotic dynamics |
| --- | --- |
| D (s) | cooperators and punishers go extinct, defectors show stable persistence |
| CP (s) | cooperators and punishers show stable persistence, defectors go extinct |
| CPD (o) | cooperators, punishers and defectors show oscillatory behaviour |
| DC (s) | defectors and cooperators show stable persistence, punishers go extinct |
| CPD (s) | cooperators, punishers, and defectors show stable persistence |
| P (s) | defectors and cooperators go extinct, punishers show stable persistence |
| PD (os) | cooperators go extinct, punishers and defectors show oscillatory behaviour |
| Os | Other dynamical regimes |

**Table 2.**
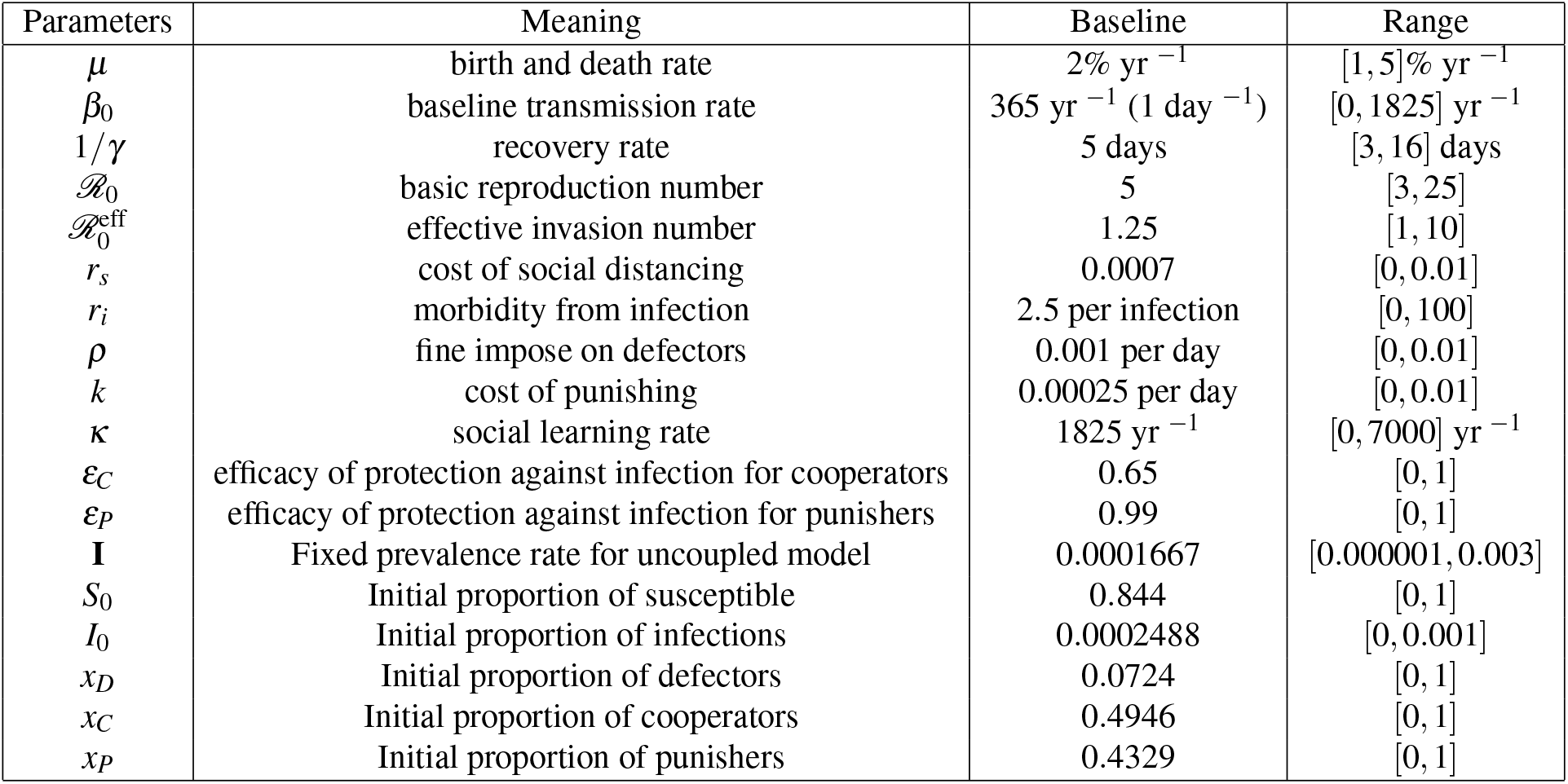
Baseline parameter values and initial conditions. Initial identified condition for stable co-existence amongst the social distancing behaviours.

| Baseline Values and Initial Conditions |  |  |  |
| --- | --- | --- | --- |
| Parameters | Meaning | Baseline | Range |
| $\mu$ | birth and death rate | 2% yr<sup>-1</sup> | [1, 5] % yr<sup>-1</sup> |
| $\beta_0$ | baseline transmission rate | 365 yr<sup>-1</sup> (1 day<sup>-1</sup>) | [0, 1825] yr<sup>-1</sup> |
| $1/\gamma$ | recovery rate | 5 days | [3, 16] days |
| $\mathcal{R}_0$ | basic reproduction number | 5 | [3, 25] |
| $\mathcal{R}_0^{\text{eff}}$ | effective invasion number | 1.25 | [1, 10] |
| $r_s$ | cost of social distancing | 0.0007 | [0, 0.01] |
| $r_i$ | morbidity from infection | 2.5 per infection | [0, 100] |
| $\rho$ | fine impose on defectors | 0.001 per day | [0, 0.01] |
| $k$ | cost of punishing | 0.00025 per day | [0, 0.01] |
| $\kappa$ | social learning rate | 1825 yr<sup>-1</sup> | [0, 7000] yr<sup>-1</sup> |
| $\varepsilon_C$ | efficacy of protection against infection for cooperators | 0.65 | [0, 1] |
| $\varepsilon_P$ | efficacy of protection against infection for punishers | 0.99 | [0, 1] |
| **I** | Fixed prevalence rate for uncoupled model | 0.0001667 | [0.000001, 0.003] |
| $S_0$ | Initial proportion of susceptible | 0.844 | [0, 1] |
| $I_0$ | Initial proportion of infections | 0.0002488 | [0, 0.001] |
| $x_D$ | Initial proportion of defectors | 0.0724 | [0, 1] |
| $x_C$ | Initial proportion of cooperators | 0.4946 | [0, 1] |
| $x_P$ | Initial proportion of punishers | 0.4329 | [0, 1] |

**Figure 2.**
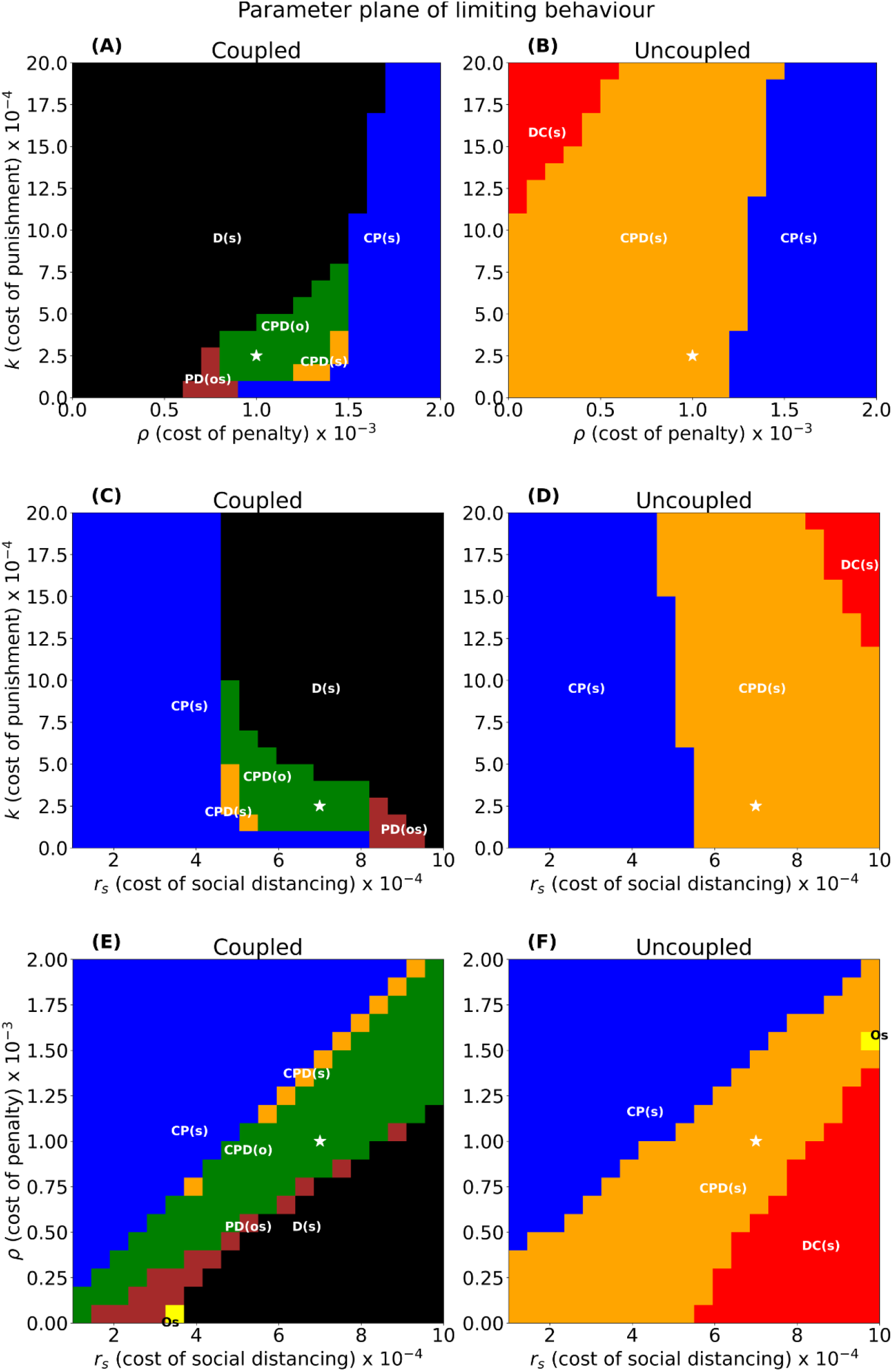
Dynamical regimes for limiting behaviour for both coupled (left panel) and uncoupled (right panel) models: parameter planes (**(A),(B)**) for cost of punishment (*k*) and cost of penalty (*ρ*), (**(C),(D)**) the cost of punishment (*k*) and cost of social distancing (*r*_*s*_), (**(E),(F)**) the cost of penalty (*ρ*) and cost of social distancing (*r*_*s*_) while all other parameter values are held at baseline in Table 2. The star represents the baseline values, and full description of legend is located in Table 1. Abbreviations for parameter regimes appear in Table 1.

### Differences between coupled and uncoupled model

Despite the broad similarities in the response of model dynamics to cost parameters, the coupled and uncoupled models show important differences in other respects. For example, in the uncoupled model, the punisher and–especially–the cooperator strategies are more robust and more likely to persist, even in the presence of defectors (Figures 2 and 3B, D, F versus A, C, E). Another difference is that the contrast between the states of adjacent regimes is often stronger in the coupled model. For instance, a reduction in the cost of social distancing can result in a tipping point from all-defect to a stable cooperate-punish state in the coupled model, but the same parameter change in the uncoupled model causes a more gradual change in the composition of strategies at equilibrium (Figure 2D versus C). Finally, the punisher strategy may persist in the coupled model through limit cycles, but no limit cycles occur in the uncoupled model for any of the parameter values tested.

**Figure 3.**
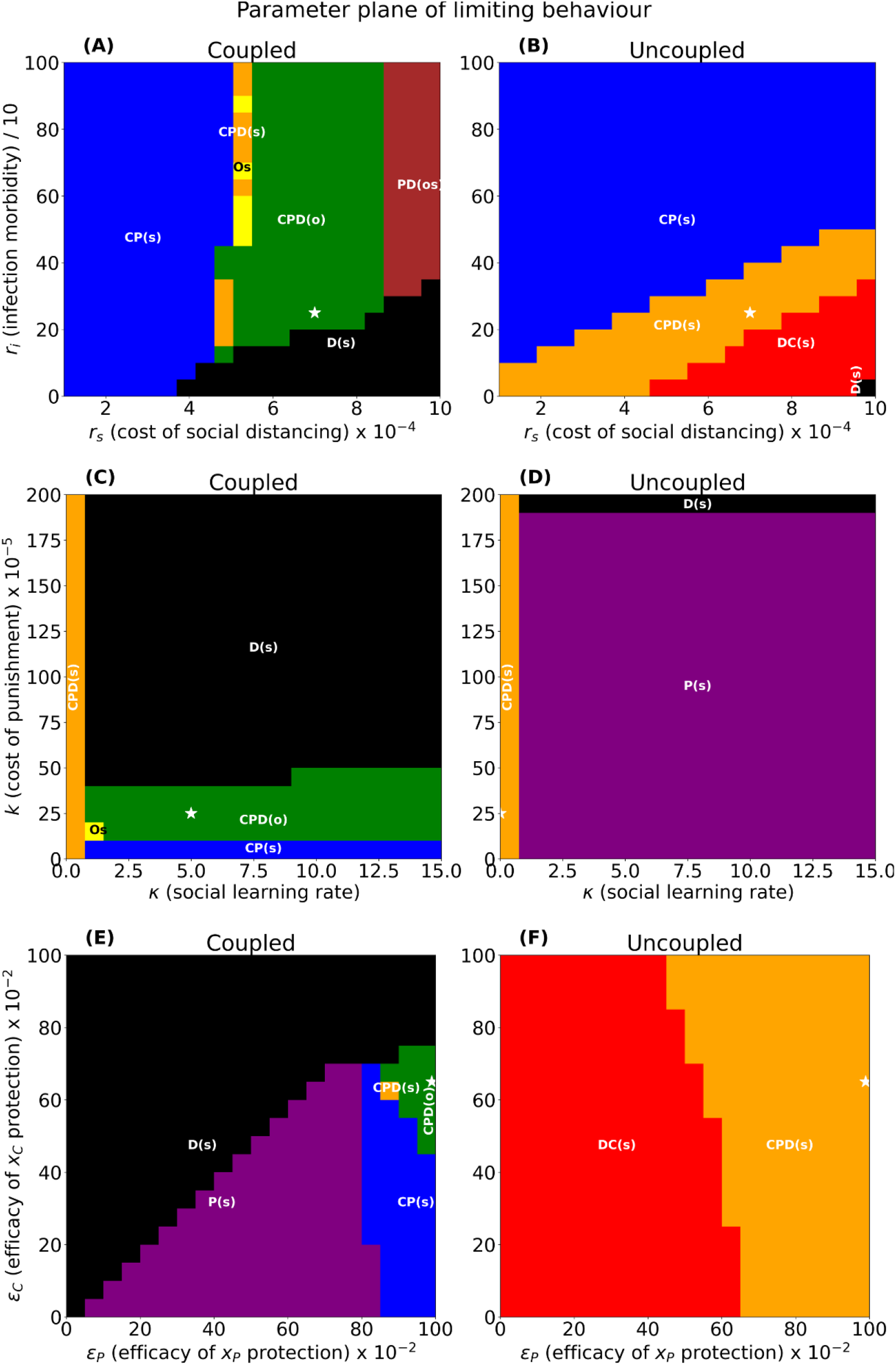
Dynamical regimes for limiting behaviour for both coupled (left panel) and uncoupled (right panel) models: parameter planes (**(G),(H)**) for morbidity from infection (*r*_*i*_) and cost of social distancing (*r*_*s*_), (**(I),(J)**) the cost of punishment (k)and social learning rate (*κ*), (**(K),(L)**) strength of protection to cooperators (*ε*_*C*_) and strength of protection to punishers (*ε*_*P*_) while all other parameter values are held at baseline in Table 2. The star represents the baseline values, and full descriptio**7**n**/1**o**4**f legend is located in Table 1. Abbreviations for parameter regimes appear in Table 1.

### Role of other parameters

A higher cost of infection (*r*_*i*_) tends to support both cooperator and punisher strategies, as expected (Figure 3A,B). The role of social distancing efficacy is more subtle, especially in the coupled model. In the coupled model, the efficacy only needs to be slightly larger for punishers than for cooperators in order for punishers to persist (Figure 3E). If the efficacy of social distancing for punishers is sufficiently large, it actually supports the persistence of cooperators, too. In contrast, for higher values of the efficacy of social distancing for cooperators, we see not an increase in cooperation but rather a collapse of both cooperators and punishers, who are replaced by the defector strategy.

In the uncoupled model, demographic and epidemiological parameters do not play a role since the prevalence is fixed. But in the coupled model, an increase in the transmission rate (*β*) supports the punisher strategy, sometimes even to the exclusion of any other strategy (Figure 4A,C,E). Conversely, an increase in the recovery rate (*γ*) supports defection, as it results in a faster elimination of the virus. An increase in the demographic turnover (births/deaths *µ*) supports the punisher strategy if the transmission rate is high enough or the recovery rate is low enough.

**Figure 4.**
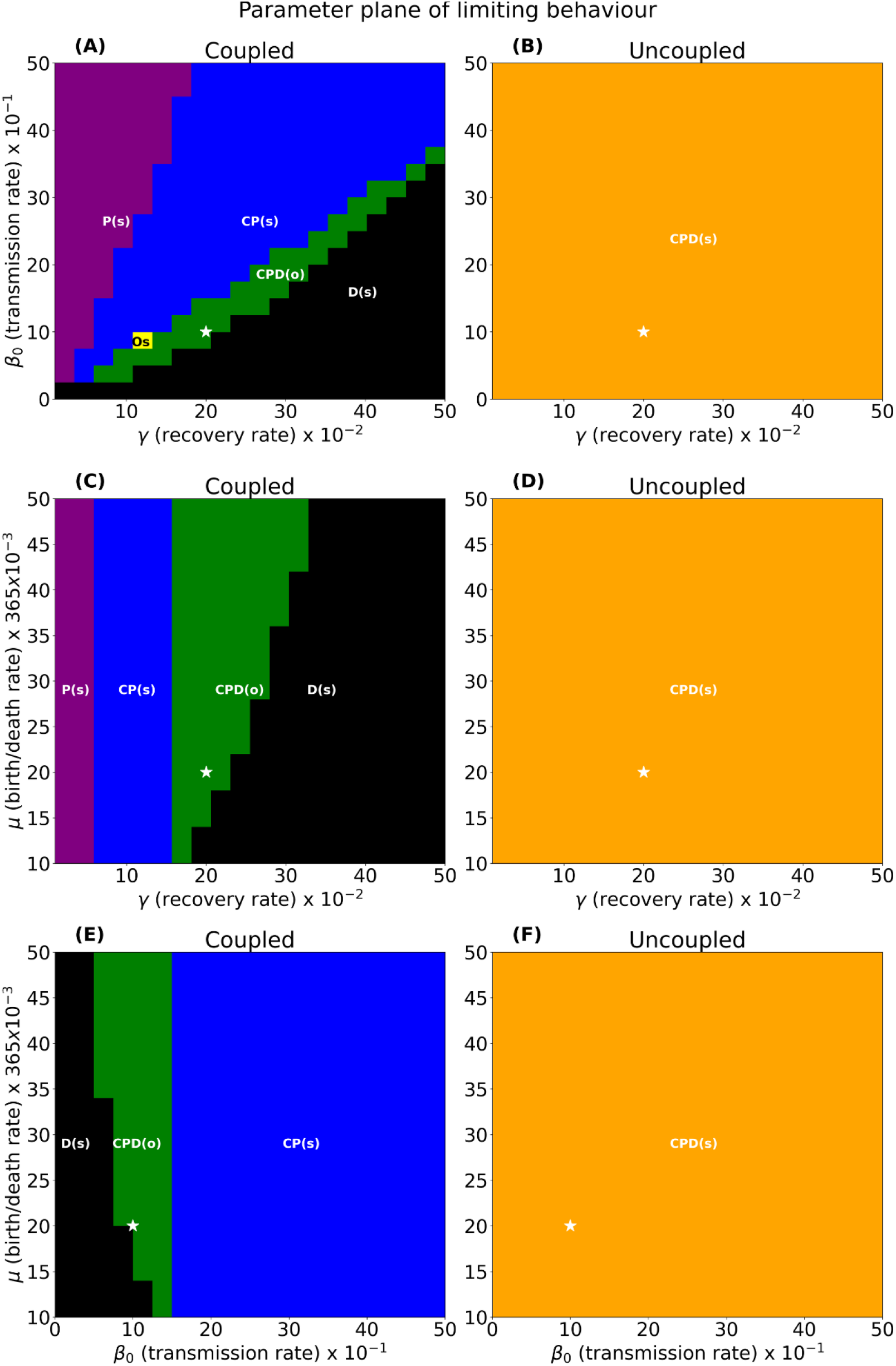
Dynamical regimes for limiting behaviour for both coupled (left panel) and uncoupled (right panel) models: parameter planes (**(M),(N)**) for baseline transmission rate (*β*_0_) and recovery rate (*γ*), (**(O),(P)**) the birth/death rate (*µ*) and recovery rate (*γ*), (**(Q),(R)**) birth/death rate (*µ*) and baseline transmission rate (*β*_0_) while all other parameter values are held at baseline in Table 2. The star represents the baseline values, and full description of legend is located in Table 1. Abbreviat**8**io**/1**n**4**s for parameter regimes appear in Table 1.

## Discussion

Social norms can be a powerful mechanism for sustaining public goods in the absence of top-down interventions from institutions in many systems^1^, and they also sustain vaccination coverage in many populations^54^. Most existing mathematical models of social norms assume pre-existing social norms, but here we modelled the endogenous establishment and persistence of norms and what social and epidemiological conditions influence their persistence. Our results show that no extra compliance system is needed to maintain the norm; rather, altruistic punishment^12,17^ is sufficient to enforce compliance and cooperation, even though punishers must pay an additional cost to enforce penalties against defectors.

The injunctive social norms^26^ of coupled behaviour-disease models work through a majority-enforcing mechanism^50^, which is also used in other types of models of norm evolution^21^. In such cases, either a pro-mitigation or anti-mitigation norm might prevail, depending on which strategy is more numerous in the starting state of the population^50^. In our model, the effect is similar even if the mechanism involves more strategies: as punishers become more common, the number of defectors declines, which in turn reduces the costs that punishers must pay to apply penalties–a positive feedback loop. Also, we do not allow for a punisher strategy that punishes social distancing.

Because this approach to norms relies upon enforcing the most common behaviour, it also raises an important distinction between norms, cooperation, and socially optimal states^55,56^. In our model, the punishment of defectors corresponds to supporting behaviours that reduce infectious disease transmission. For any sufficiently harmful infectious disease, this behaviour will improve the average population health and thus will correspond to a socially optimal outcome, similar to what might be expected for a pure cooperator strategy. However, promoting cooperation *per se* does not always generate socially optimal states^56^. For instance, the decision of villagers in Eyam, England, during the plague outbreak from 1665 to 1666, was a voluntary act of social distancing that can be interpreted as a cooperative strategy reinforced by a short-lived social norm, although it may not have actually improved health outcomes in the population at large^57^.

In addition, there are other mechanisms by which norms can emerge in populations, aside from altruistic punishment^58^. For instance, descriptive norms are based simply on observing and copying what is a commonly practiced behaviour in a group^26^. Similarly, it’s possible that a generalized tendency to follow social norms has evolved in humans, aside from being motivated by specific conditions where the meaning of defection is agreed upon and the socially optimal outcome is clear to everyone^49^. Hence, altruistic punishment is not the only way that social norms in support of social distancing might emerge.

We acknowledge some limitations of our approach. Notably, we assumed that the efficacy of social distancing is higher for punishers, since an easily accessible means of punishing is to ostracize those who do not practice social distancing. It is reasonable to assume that punishing those who do not socially distance is positively associated with a higher personal efficacy of social distancing, but it does alter the cost-benefit balance of the punisher strategy. Despite our assumption that punishers benefited from a higher efficacy of social distancing, the efficacy only had to be slightly higher for the punishers than for the cooperators in order for the punisher strategy to persist (^**?**^E). We also assumed a homogeneous population. Heterogeneity can influence the predictions of coupled human-environmental models^59,60^. Some important factors that we did not address include cultural differences in response to social processes^61^; the impact of economic factors in determining the cost of social distancing and how those vary between lower- and higher-income countries^62^; the possibility that distancing costs may change over time^62^; and the role of other cultural and religious influences^63,64^. Taking these limitations into consideration and making appropriate adjustments to the model can be undertaken in future work. We also did not address the intervention of vaccination and its associated social norms^65^, although vaccination is a major intervention against many infectious diseases with a Susceptible-Infectious-Recovered natural history as we studied here^66^. This is another promising area for future research. In conclusion, we have shown how a strategy of punishing those who do not practice social distancing can persist in a population, even when punishing incurs a cost on the punishers. Altruistic cooperation induced by altruistic punishment for social distancing is possible when a section of the population is willing to bear a cost to themselves to punish others to enforce cooperation. This punishment-induced cooperation can be sustained as it becomes a social norm, that is, a socially acceptable public good. The infectious disease can consequently be mitigated or eliminated from the population.

## Methods

### Infectious disease model

Disease transmission is modelled with a compartmental (SIR) model^67^. Birth and death are included since we are interested in the long-term endogenous dynamics of norms. The disease causing pathogen invades the susceptible (*S*) population and infects an individual; the infected individual becomes infectious (*I*) and is capable of infecting others. At this stage, the individual can die from the disease or recover to a state of immunity. Individuals in any compartment can also die of other causes. Note here that we do not consider the compartment of the social distancing group population, as is commonly used in compartmental models involving disease mitigation studies, but we model social distancing as a social behaviour where decisions can evolve over time.

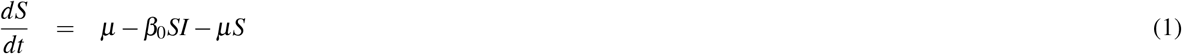

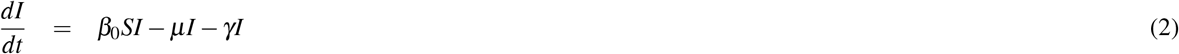

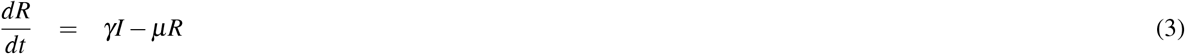

The proportion of susceptible individuals is represented by (*S*), the proportion of infected individuals as (*I*), the proportion of recovered individuals as (*R*), *µ* represents the birth and death rates (birth rate = death rate), *β*_0_ is the baseline transmission rate in the absence of mitigation strategies, and *γ* is the recovery rate.

### Social distancing strategies

Social distancing is modelled by considering three behavioural strategies that the population can adopt – a non mitigating strategy (defectors) and two mitigating strategies (cooperators and punishers). Punishers are like cooperators, but they also pay a cost to impose a penalty against defectors. Let the proportion of individuals defecting be *x*_*D*_, cooperating be *x*_*C*_, and punishers be *x*_*P*_. Infected individuals perceive a risk of infection that is simply proportional to the current prevalence *I*, and if infected, they perceive a probability *r*_*i*_ of suffering significant morbidity. Defectors suffer a penalty *ρ* imposed on them by each punisher. Both cooperators and punishers bear the cost *r*_*s*_ of social distancing since it costs to distance oneself from society. (We only consider costs to individuals due to their choices, not the social costs of a broader shutdown, which we do not model here.) Social ostracism is considered a costly punishment, so punishers pay a cost *k* to punish each defector. The efficacy of social distancing in reducing the infection risk for cooperators (punishers) is *ε*_*C*_ (*ε*_*P*_), respectively, and defectors (*ε*_*D*_ = 0) do not reduce their infection risk at all since they do not practice social distancing. Thus, the payoffs for the strategies are:

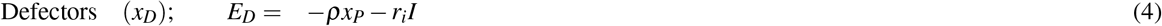

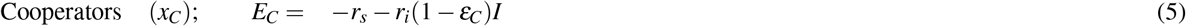

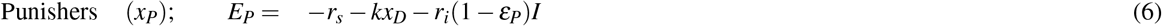

### Imitation Dynamics

The social distancing strategies of individuals evolve over time, allowing an individual to adjust their strategy by adopting a different method through various approaches such as imitating higher utility strategies, following a specific stochastic process (e.g., frequency dependent Moran process), or using the proportional imitation rule. In this work, we use the proportional imitation rule from evolutionary game theory, modelled as the replicator dynamic equation^68^. An individual is selected from the population and given the chance to change their behaviour. The selected individual is allowed to sample another individual randomly and is offered the opportunity to adopt their strategy with a certain probability.

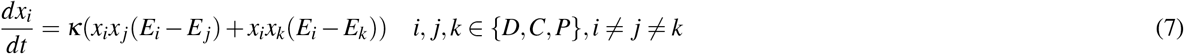

where *x*_*i*_*x* _*j*_, *x*_*i*_*x*_*k*_ are the probabilities of randomly and independently sampling the individual from *x* _*j*_ or *x*_*k*_, and then potentially imitating the individual’s behaviour type *i ∈ {D,C, P}*. This leads to the following equations for imitation dynamics in the social distancing game.

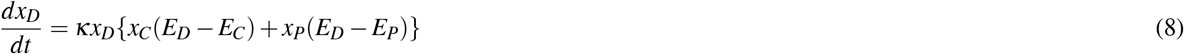

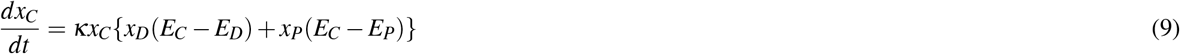

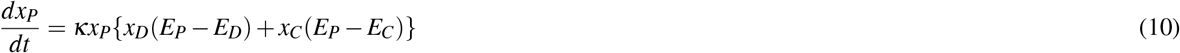

### Coupled behaviour-disease model

The change in the strategy composition of a population during a disease outbreak always influences the dynamics of the disease outcome as a feedback mechanism. A mitigating strategy such as social distancing will cause the case incidence to reduce to an acceptable threshold level – *R*_0_ *<* 1 (endemic state) provided that a significant portion of the population adheres to the precautionary contact measures. On the other hand, a non-mitigating strategy such as defecting will not change the transmission rate of the disease, potentially causing it to persist within the population. As a result of this feedback mechanism between social dynamics and disease dynamics, we can couple the SIR model with the social distancing behaviour model to obtain our coupled behaviour-disease model. One method of coupling the two models is by using the contact precaution approach, where the feedback mechanism influences the transmission rate (*β*) directly, thus *β* = *β*_0_(*x*_*D*_ + *x*_*C*_(1 *−ε*_*C*_) + *x*_*P*_(1 *−ε*_*P*_)) giving us the following model equations;

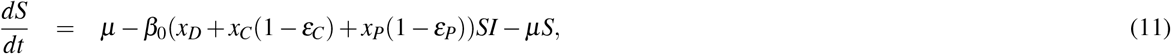

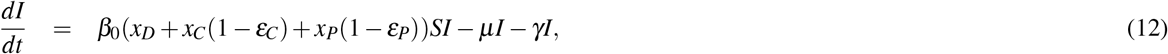

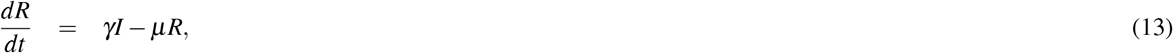

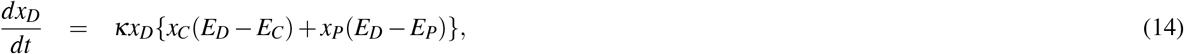

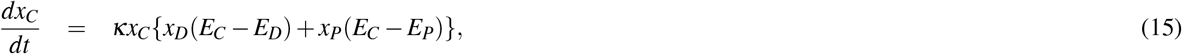

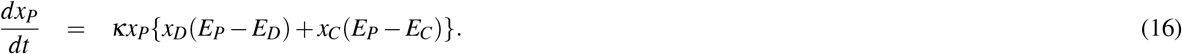

Since *S* + *R* + *I* = 1*⇒ R* = 1*− S− I* and *x*_*D*_ + *x*_*C*_ + *x*_*P*_ = 1 ⇒ *x*_*C*_ = 1 *− x*_*D*_*− x*_*P*_, we obtain the model equations for contact precautions. We include case importation for realism and to help stabilize numerics near the boundary (*η* = 10^*−*7^):

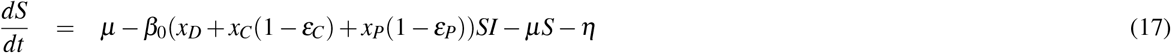

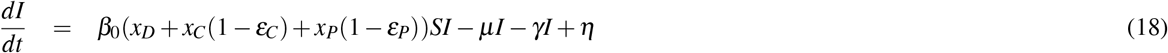

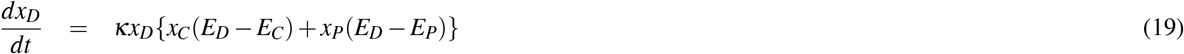

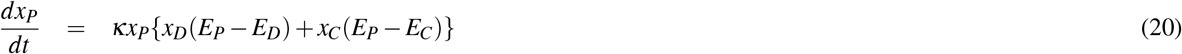

which gives a four dimensional model instead of six. We can obtain the endemic equilibrium and establish conditions under which stable co-existence is guaranteed. We do so by considering 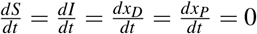 Let Θ = (*x*_*D*_ + *x*_*C*_(1 *− ε*_*C*_) + *x P* (1 *− ε P*)); then we can obtain the endemic equilibrium (*I*_0_ *>* 0 and *ℛ*_0_ *>* 1) as 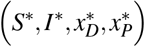 = 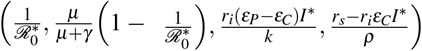 where 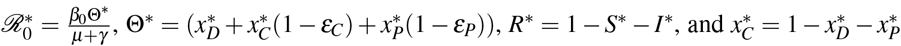 To ensure we have non-zero initial conditions, we set the following necessary conditions: 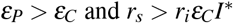

### Uncoupled behaviour-disease model

In the previous sections, we discussed the case where the payoffs vary with the infection prevalence; that is, a feedback mechanism between the disease system and the social distancing behaviour system. In such a model, the population has access to resources such as information to help inform their imitation of others. We consider a control scenario where the infection prevalence (and thus payoff) is fixed at *I*(*t*) = **I**, resulting in a fixed payoff matrix. The system remains a behaviour-disease model, but the feedback mechanism becomes one-way; that is, the social distancing behaviour influences the disease dynamics, but there is no corresponding feedback from the current prevalence level on the behaviour strategies. Thus, the expected payoff for each strategy becomes:

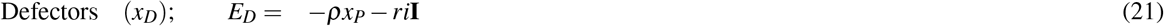

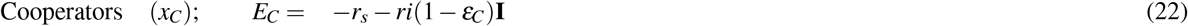

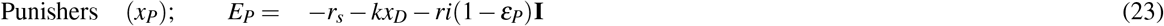

### Stability and Limiting Behaviours

We estimate the baseline values and initial conditions to obtain stable co-existence between the defectors and punishers (Table 2). Stability is essentially the case where a small perturbation of the initial conditions does not cause significant changes in the abundance of behaviour of the strategies (local asymptotic stability), while co-existence is maintained at non zero proportions for *t > T →* ∞. As such, we are interested in either stable nonzero or limit cycle behaviours. We are also interested in other possible conditions that guarantee stable co-existence. We employ parameter planes with the baseline values and initial conditions for this exercise. With this, we show other possible limiting behaviours in the parameter plane that require definition, resulting in a classification of limiting behaviours. We identify three main behaviours: extinction (when the proportion of a behaviour type is less than 0.1%), stable nonzero (when the proportion of a behaviour type remains at a constant rate greater than 0.1% for *t >>* 1), and oscillatory behaviours. The oscillatory behaviours consist of limit cycles (when the trajectory or dynamics exhibit periodic oscillations of possibly equal phase and amplitude), chaotic (when the trajectory shows oscillations that are bounded but have varying levels of phase and amplitude), quasi periodic, and other more complex dynamics (oscillations that are unbounded with increasing amplitude or appear to be fixed points but with small levels of amplitude and phase). We identify the limiting behaviour by simulating the model for each varying parameter pair for at least 2000 years for the coupled model and 5000 years for the uncoupled model. We then consider the last 400 years and determine the behaviour based on the three classes of such time series in Python. These limiting behaviours are used to create the dynamical regimes in Table 1.

### Parameters Values and Initial Conditions

The complete system is evaluated numerically, where model parameters are assessed to obtain stable fixed states or limit cycles in which co-existence is maintained as punishers and defectors remain in the population. The baseline values are then obtained accordingly using the assumed ranges for the parameters and initial conditions, as shown in Table 2.

## Data Availability

All data produced in the present study are available upon reasonable request to the authors

## Declaration of Competing interest

The authors declare that they have no known competing financial interests or personal relationships that could have appeared to influence the work reported in this paper.

### Authors Contribution

CTB conceived the model, and SF established the results. SF draughted the manuscript, and CTB reviewed and edited it. Both authors read and approved the final version of the manuscript. SF accessed and verified the data used in the study and had final responsibility for the decision to submit for publication.

## Acknowledgement

This work was supported by a Discovery Grant to CTB from the Natural Sciences and Engineering Council of Canada (NSERC) [grant number RGPIN-2025-04274]. The funder of the study had no role in study design, data collection, data analysis, data interpretation, writing of the paper, or the decision to submit.

